# A data-adaptive method for investigating effect heterogeneity with high-dimensional covariates in Mendelian randomization

**DOI:** 10.1101/2023.10.28.23297706

**Authors:** Haodong Tian, Brian D. M. Tom, Stephen Burgess

## Abstract

Mendelian randomization is a popular method for causal inference with observational data that uses genetic variants as instrumental variables. Similarly to a randomized trial, a standard Mendelian randomization analysis estimates the population-averaged effect of an exposure on an outcome. Dividing the population into subgroups can reveal effect heterogeneity to inform who would most benefit from intervention on the exposure. However, as covariates are measured post-”randomization”, naive stratification typically induces collider bias in stratum-specific estimates. We extend a previously proposed stratification method (the “doubly-ranked method”) to form strata based on a single covariate, and introduce a data-adaptive random forest method to calculate stratum-specific estimates that are robust to collider bias based on a high-dimensional covariate set. We also propose measures to assess heterogeneity between stratum-specific estimates (to understand whether estimates are more variable than expected due to chance alone) and variable importance (to identify the key drivers of effect heterogeneity). We show that the effect of body mass index (BMI) on lung function is heterogeneous, depending most strongly on hip circumference and weight. While for most individuals, the predicted effect of increasing BMI on lung function is negative, it is positive for some individuals and strongly negative for others.

## Introduction

Mendelian randomization uses genetic variants as instrumental variables to investigate the causal effect of a modifiable exposure on a health outcome [18]. Randomness in the allocation of genetic variants from parent to offspring can be exploited in a natural experiment, analogous to a randomized controlled trial [43]. Under Mendel’s laws of segregation and independent assortment, between-sibling genetic associations should be unaffected by confounding, and so genetic variants should only be associated with traits that they affect. Under the further assumption that any causal pathway from the genetic variants to the outcome passes via an intermediate exposure, a genetic association with the outcome is indicative of a causal effect of the exposure on the outcome [19]. Empirical investigations have suggested that genetic variants behave similarly to randomization at a population level for large well-mixed populations [44, 49], meaning that Mendelian randomization can be used to make reliable causal inferences even in population-based datasets. Randomized trials typically estimate an average causal effect, representing the effect of varying the exposure averaged across all individuals in the population [1]. However, it may be that the effect of the exposure on the outcome differs amongst individuals in the population. This is often addressed by performing stratified analyses: dividing the population into subgroups and estimating separate effects in each subgroup [53]. However, care is required, as stratification on a variable that is a common effect of two variables (known as a collider) leads to a correlation between those two variables within the strata, even if they are uncorrelated in the population as a whole [16]. Hence, while random allocation in a trial should be independent of all potential competing risk factors in the overall trial population measured at baseline, stratification on a variable that is an effect of randomization can lead to associations with competing risk factors, and hence to bias in subgroup estimates (known as collider bias). In trials, a sharp division is made between stratification on a pre-randomization or baseline covariate, versus stratification on a post-randomization covariate; the latter have been called “improper” subgroup analyses, as they are at risk of collider bias [55]. However, in Mendelian randomization, as the “randomization” event occurs at an individual’s conception, all covariates (except for those not subject to the effects of genetic variation, such as age, sex, and measures of ancestry) are post-randomization covariates.

Previous methodological investigations have shown that stratification on a covariate can lead to bias in Mendelian randomization estimates [14, 23], and potentially misleading results due to stratification have been observed in applied analyses [10]. Two approaches have been proposed for stratification that avoid collider bias: the residual method [17] and the doubly-ranked method [50]. The residual method first calculates the residual from regression of the covariate on the genetic variants, and stratifies based on the residual values of the covariate. The doubly-ranked method first divides the population into pre-strata based on levels of the genetic variants, and then forms strata by picking individuals from each pre-stratum based on levels of the exposure. The residual method assumes that the effect of the genetic variants on the exposure is linear and homogeneous in the population [41], whereas the doubly-ranked method makes a weaker ‘rank-preserving assumption’: that the genetic variants do not affect the ranking of participants according to their levels of the exposure. In the context of non-linear Mendelian randomization, where we form strata based on levels of the exposure, the doubly-ranked method has been shown to be less sensitive to variation in the effect of the genetic variants on the exposure compared to the residual method [11].

In this paper, we extend the doubly-ranked method to consider stratification on a covariate, and introduce a data-adaptive random forest method that allows feasible and efficient investigation of stratum-specific Mendelian randomization estimates based on a high-dimensional set of covariates. We demonstrate the utility of this method in a simulation study, and an applied analysis into the effect of body mass index (BMI) on lung function. We show that the effect of BMI on lung function varies strongly, with negative estimates for most individuals in the population, but positive estimates for others. We conclude by discussing the relevance of these investigations for the design of clinical trials. The code for implementing the effect heterogeneity analysis in MR is available at https://github.com/HDTian/RFQT.

## Results

### Random forest of Q trees method

We assume a single dataset with individual-level data on an exposure, an outcome, a genetic instrument, and a high-dimensional set of candidate covariates, some of which may be effect modifiers. A Q tree is formed by recursively dividing the population into groups (Figure 1). At each node, we form two strata based on each covariate in turn, calculate stratum-specific Mendelian randomization estimates, and choose the covariate that gives rise to the greatest value of the Q statistic, a measure of heterogeneity amongst the stratum-specific estimates. We then divide into two nodes based on the stratification value of that covariate. We stop when any one of the stopping rules is met. We then calculate Mendelian randomization estimates in the terminal nodes using the ratio method. The causal interpretation of the Mendelian randomization estimate is the total effect of the exposure on the outcome at the values of the covariates that we stratify on, averaged across individuals in that stratum.

**Figure 1:**
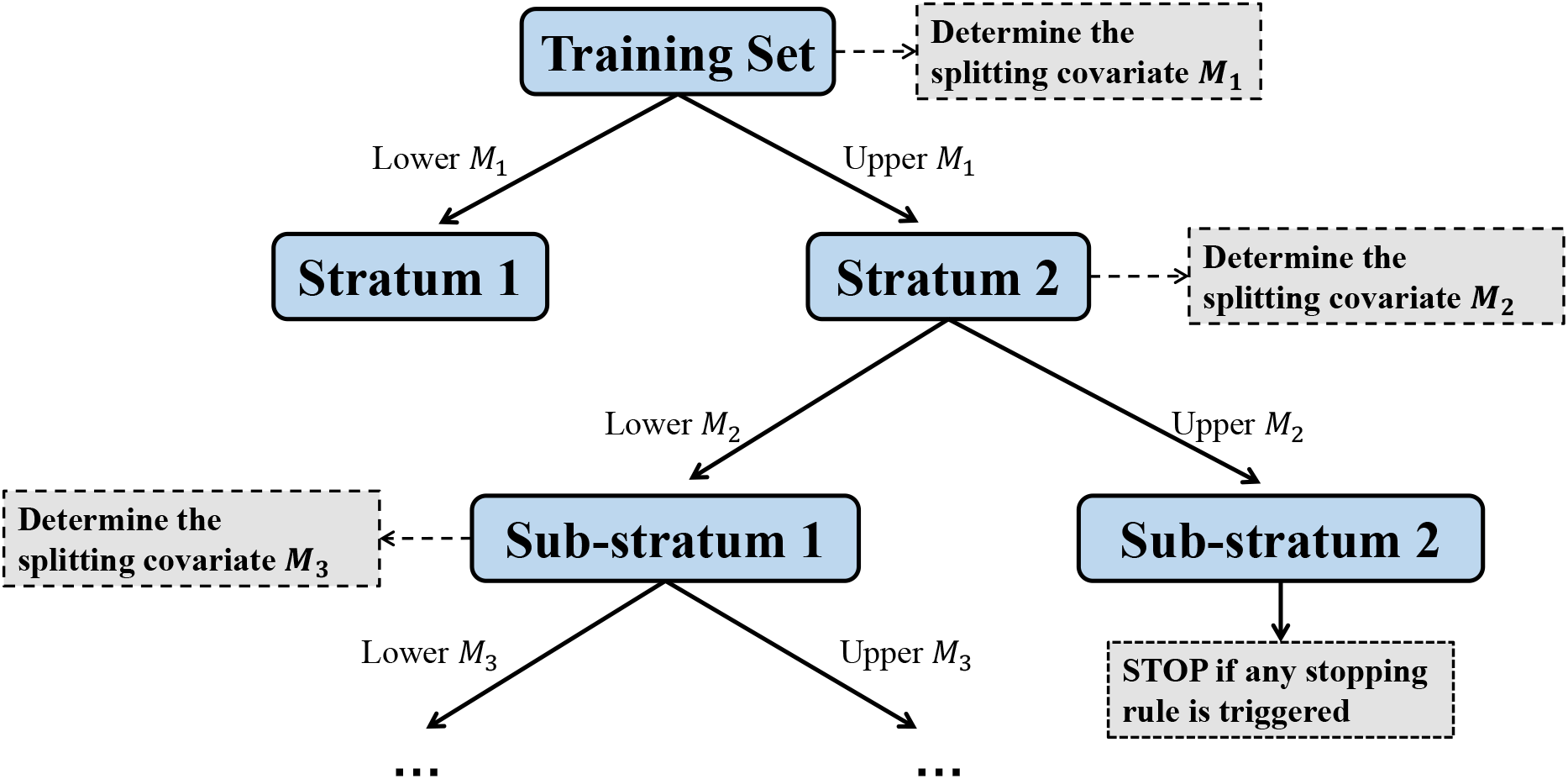
Schematic diagram illustrating the process of constructing a single Q tree. At each node, the covariate with the greatest value of the Q statistic is selected. The same covariate is allowed to be selected multiple times at downstream nodes. In each split, two nodes are formed according to the values of the covariate selected. The lower/upper *M* refers to the residual values in the lower/upper quantile region when using the residual method, or the lower/upper *M* samples in each pre-stratum when using the doubly-ranked method. The node will stop splitting when it meets any one of the stopping rules: (i) the split will cause the node size to be less than 1000, (ii) the Q-statistic value of the chosen covariate is less than 3.84 (the 95th percentile of a chi-squared distribution with one degree of freedom), or (iii) the maximum tree depth for the node is larger than 5.

To reduce the variance of the estimator, increase stability, and smooth decision boundaries [9, 39], we aggregate information from multiple de-correlated trees using a random forest of Q trees method (Figure 2). We divide the original dataset into a training subset and a testing subset. We then take multiple bootstrap samples of the training subset. For each bootstrap sample, we calculate a Q tree as described above, except that we only consider 40% of the candidate covariates at each node; this reduces correlation between separate trees. We then obtain estimates for each individual in the training and testing subsets for each tree, and average these estimates across the trees. We also calculate variable importance measures for each covariate on out-of-bag individuals (i.e. those in the training subset who were not selected into the bootstrap sample) averaged across trees. and perform a permutation test on estimates for individuals in the training subset to assess whether variability in estimates is greater than would be expected due to chance alone. Further details are provided in the Methods.

**Figure 2:**
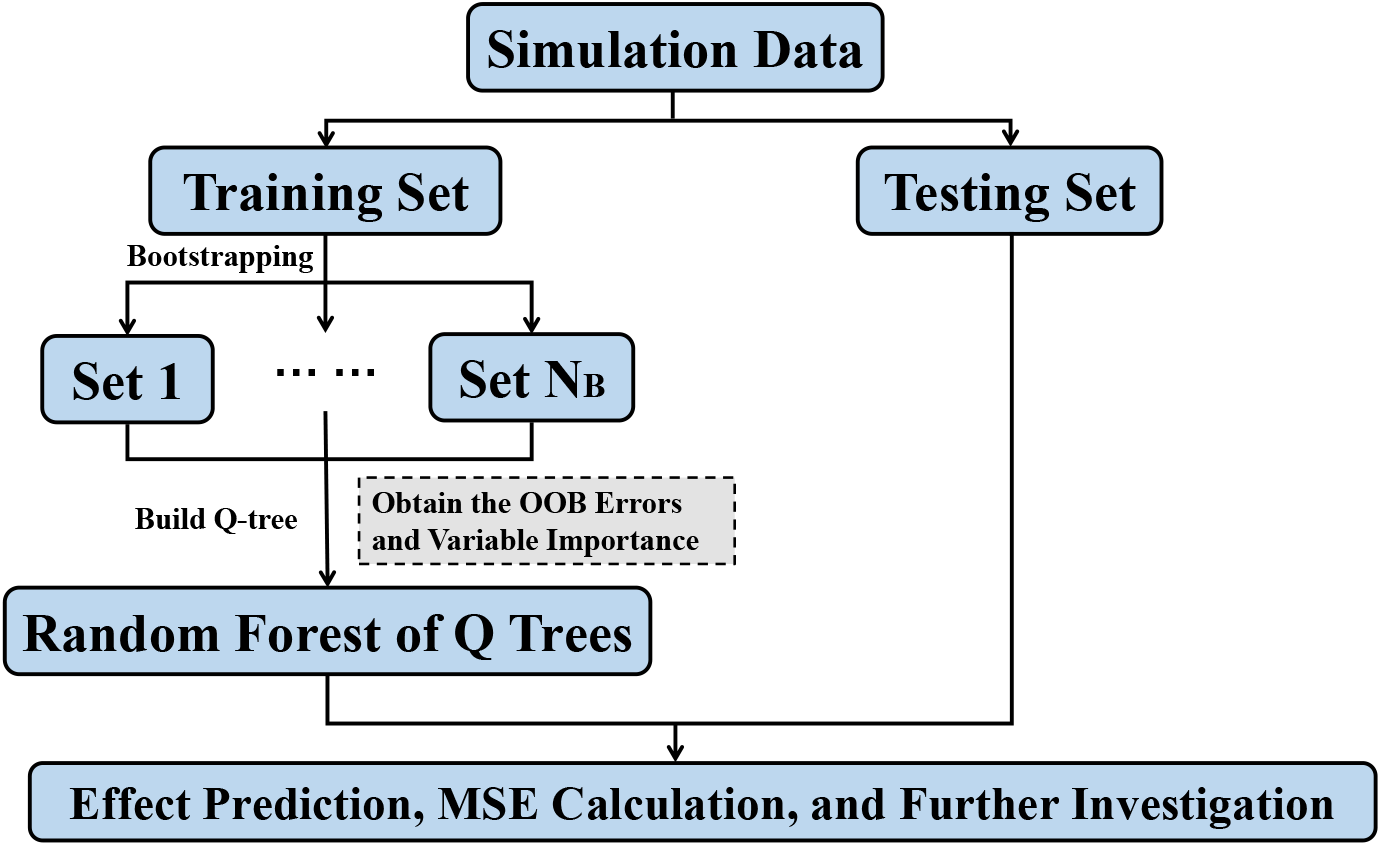
Schematic diagram illustrating the process of constructing a random forest of Q trees. OOB: Out-of-Bag. *N*_*B*_: The number of bootstrap samples and Q trees.

### Simulation study

We perform a simulation study to assess the performance of our methods. We consider three scenarios in which the true causal effect of the exposure on the outcome varies in the population (see Methods). In Scenario A, the effect varies based on covariates that are not colliders. In Scenario B, the effect varies based on covariates, some of which are colliders. In Scenario C, the effect varies based on covariates that are colliders in a complex and heterogeneous way. We compare three methods for forming strata: naive stratification on covariates, the residual method, and the doubly-ranked method, and two methods for constructing estimates: a single Q tree and the random forest of Q trees approach. We also compare results with no stratification. In total, seven methods are compared across the three scenarios. For each method, we calculate the mean squared error (MSE) of the individual-level estimates with varying levels of effect heterogeneity. The effect estimates are compared with effect values calculated from the data-generating model. We also calculate variable importance measures, and assess whether the true effect modifiers are correctly identified. We calculate variable importance measures in two ways: first, by comparing changes in MSE obtained from effects calculated using the data-generating model; and second, based on changes in the predicted effect estimates. The second approach reflects typical practice outside of a simulation setting, where the true effects are unknown.

Results in Figure 3 show that the stratification methods performed similarly in Scenario A, as the covariates are not colliders, and so are independent of the instrument. The random forest approach outperformed the single tree and no stratification approaches. In Scenario B, random forests implementing the residual and doubly-ranked stratification methods performed best, as the assumptions are satisfied for both methods. In Scenario C, the random forest implementing the doubly-ranked stratification method performed best at most levels of effect heterogeneity, particularly when the heterogeneity strength is strong. The doubly-ranked method with random forest correctly identified the true effect modifiers, whether variable importance measures were calculated using the true effects or not (Supplementary Figure S1). A scatterplot of the predicted effects against the true effects showed a strong correlation (Supplementary Figure S2).

**Figure 3:**
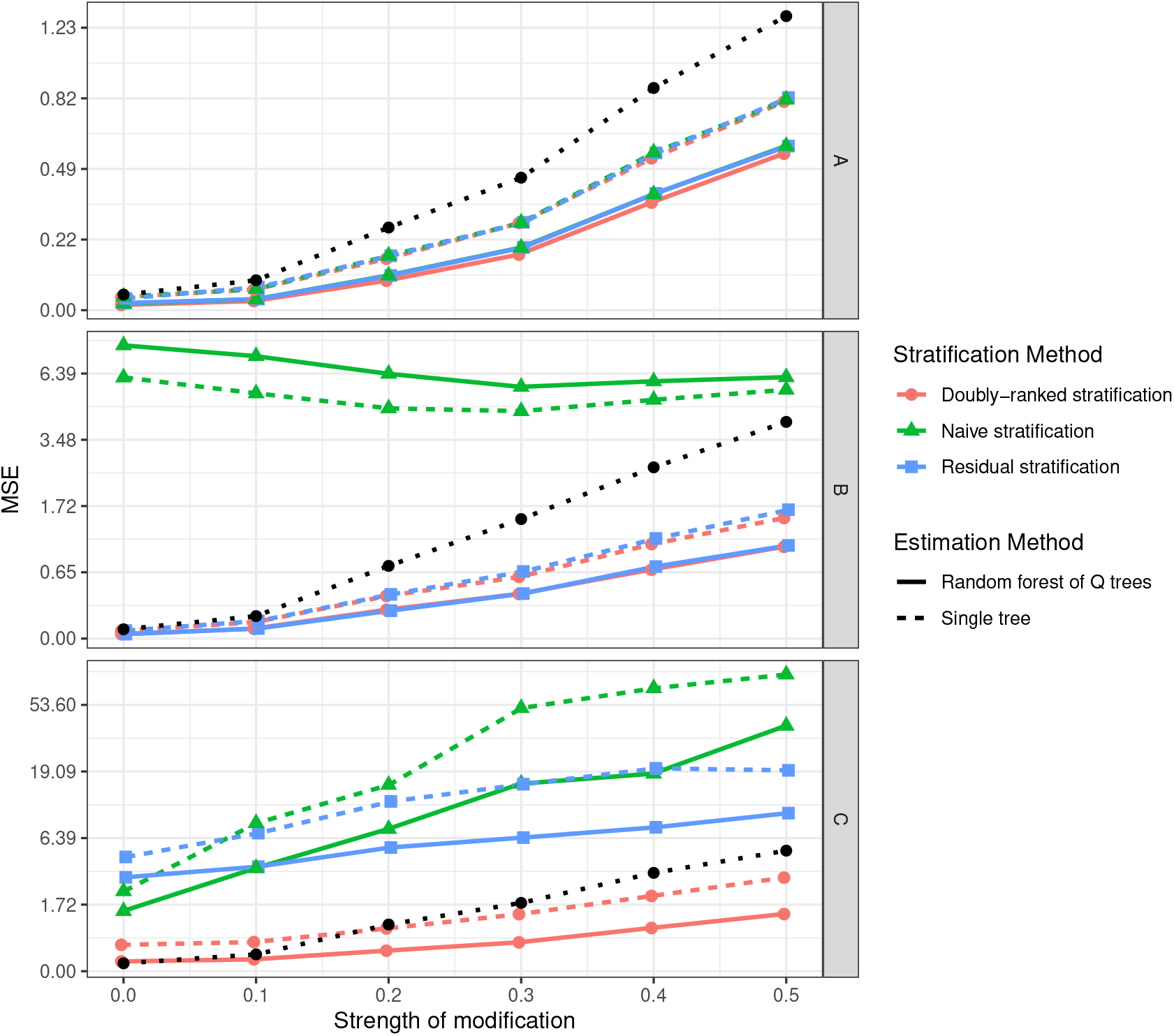
Results of the simulation study showing mean squared error (MSE) of estimates with weak modification (strength of modification = 0.0) up to strong modification (strength of modification = 0.5). Top panel: Scenario A (all effect modifiers are non-colliders); middle panel: Scenario B (some effect modifiers are colliders); bottom panel: Scenario C (effect modifiers are colliders and influence the causal effect in a complex way). The black line represents results with no stratification. In each scenario, data are independently simulated 100 times, and the MSE represents the median value across simulations..

### Stratified estimates for effect of body mass index on lung function

We considered data on 167,121 unrelated male participants of European ancestries from UK Biobank, a population-based cohort study of UK residents ages 40-69 at recruitment [47], who passed quality control checks as previously described [2]. Our exposure was BMI, measured at study entry. Our outcome was forced expiratory volume in 1 second (FEV1), also measured at study entry. Individuals were allowed up to three attempts to breathe into a spirometer; the largest recorded value was taken as the measure of FEV1. We considered 28 candidate covariates, including the exposure itself as a covariate (Supplementary Table S1). Our genetic instrument was taken as a weighted score based on 94 uncorrelated genetic variants previously shown to be associated with BMI at a genome-wide level of significance (p *<* 5 *×* 10^*−*8^) in the Genetic Investigation of ANthropometric Traits (GIANT) consortium, before the inclusion of UK Biobank in the consortium [29]. The score explained around 2% of the variability in BMI in UK Biobank participants. We took two-thirds of participants as the training subset, and one-third as the testing subset, and obtained estimates using the random forest approach and the doubly-ranked stratification method, averaging over 200 Q trees for bootstrapped samples of the training set. All estimates represent change in FEV1 measured in litres per 1 kg/m^2^ increase in genetically-predicted BMI.

A histogram of the individual-participant estimates is shown as Figure 4. We see that the distribution of estimates is positively-skewed, with most individuals having a negative estimate (i.e. higher BMI reduces lung function). Some individuals have a slight positive estimate (i.e. higher BMI increases lung function), and some individuals have a more negative estimate. There was strong evidence indicating that estimates were more variable than would be expected due to chance alone (*p <* 0.001, Supplementary Figure S3).

**Figure 4:**
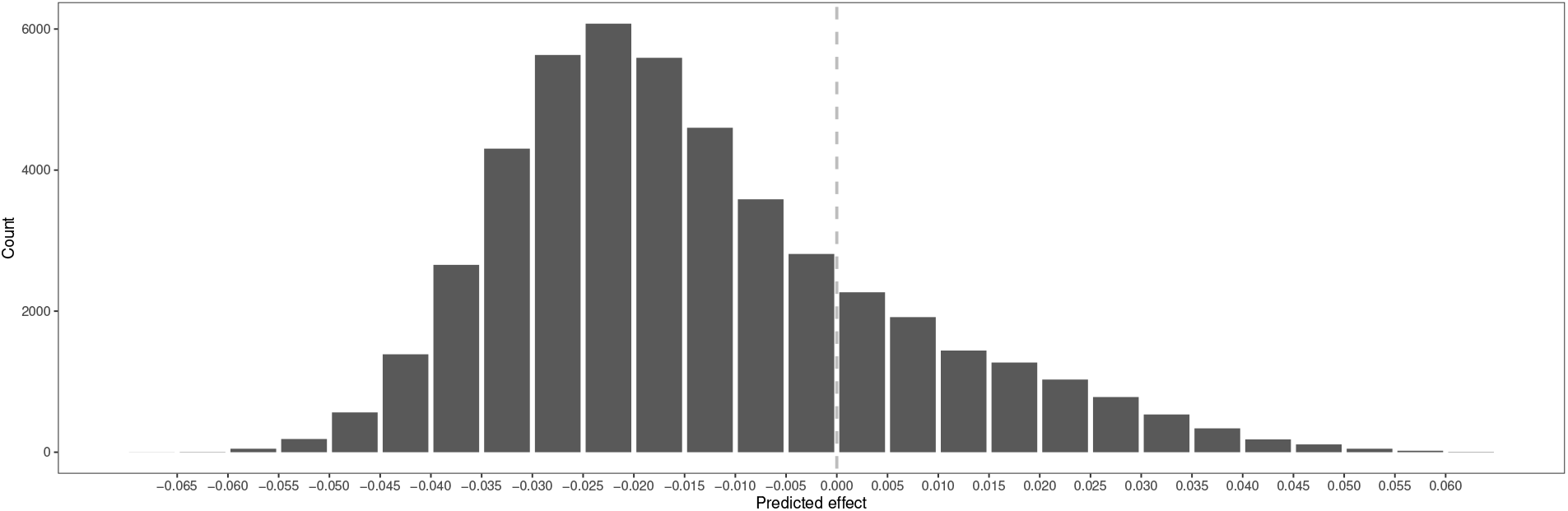
Histogram of the predicted effects of BMI on lung function from the random forest of Q trees approach using doubly-ranked stratification. Estimates represent the change in lung function (litres) per 1 *kg/m*^2^ higher genetically-predicted BMI.

Variable importance scores for the 28 covariates are shown in Supplementary Figure S4. The covariates with the highest scores were diastolic blood pressure, hip circumference, monocyte count and weight. In contrast, height was one of the lowest ranking covariates. For eight of these covariates, we divided the full dataset into tenths based on that covariate using the doubly-ranked method, and calculated stratum-specific Mendelian randomization estimates within each tenth of the population. Estimates are illustrated in Figure 5.

**Figure 5:**
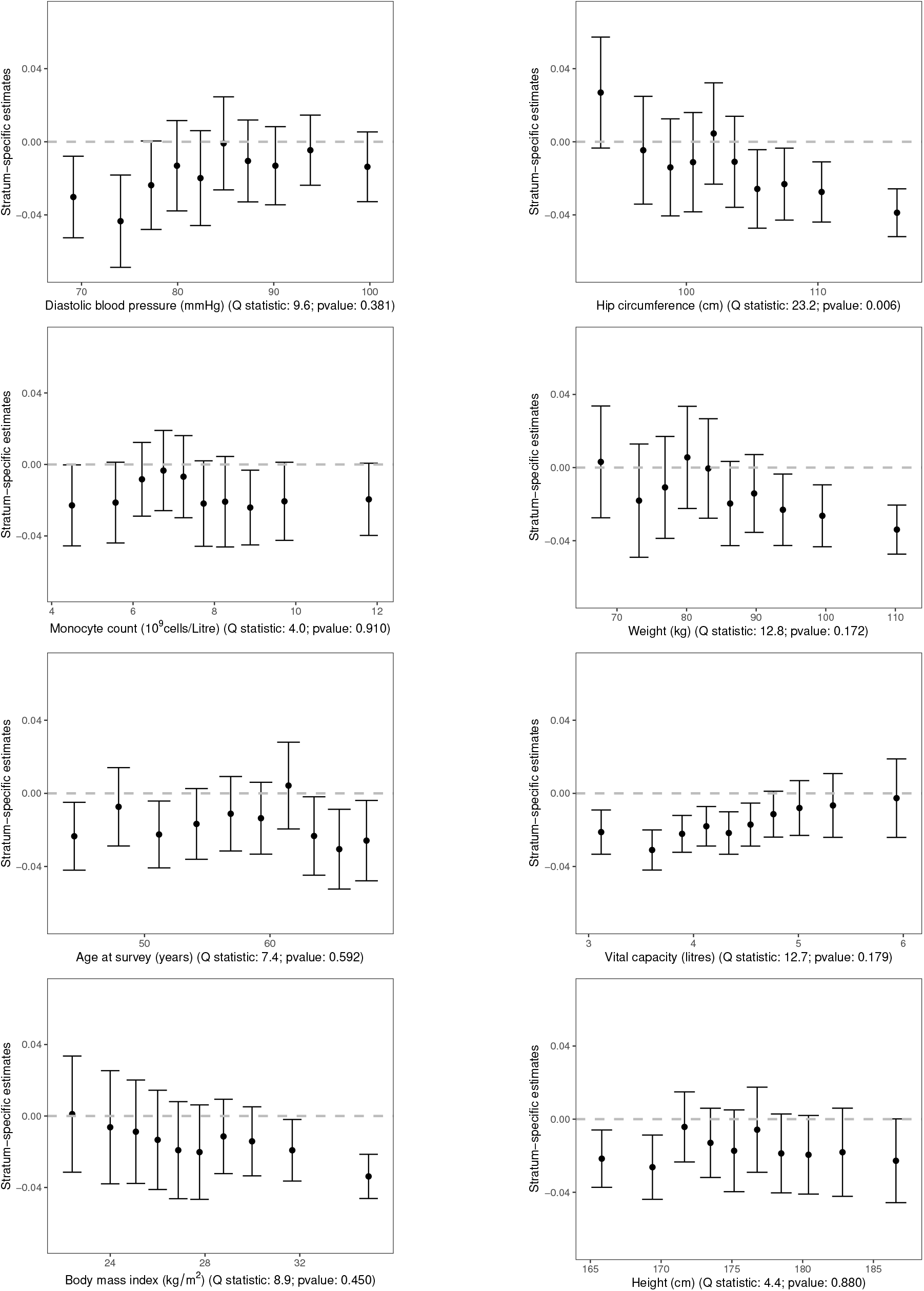
Stratum-specific estimates (error bars represent 95% confidence intervals) for the effect of BMI on lung function in deciles of the population stratified on covariates using the doubly-ranked method. The Q statistic is a measure of heterogeneity in the stratum-specific estimates.

We see that stratum-specific estimates for low values of hip circumference are compatible with the null, whereas estimates for greater values of hip circumference are negative, with some statistical evidence for heterogeneity in estimates (*p* = 0.006). This suggests that, for strata of the population with narrow hip circumference, BMI has a neutral average effect on lung function; but for strata of the population with wider hip circumference, increases in BMI lead to reduced lung function. A similar pattern was observed for weight and BMI. Trend tests indicated some evidence for a negative trend in estimates for hip circumference (*p* = 0.00001), weight (*p* = 0.002), and BMI (*p* = 0.006). In contrast, for height, there was no evidence of heterogeneity in estimates across strata, with negative point estimates in all strata (although confidence intervals overlapped the null for most strata).

More complex stratification patterns can be illustrated by plotting a decision tree. The decision tree fitting the covariate information and the predicted effects is depicted in Figure 6. The primary splitting variables for the main node are hip circumference and diastolic blood pressure, which aligns with the variable importance analysis.

**Figure 6:**
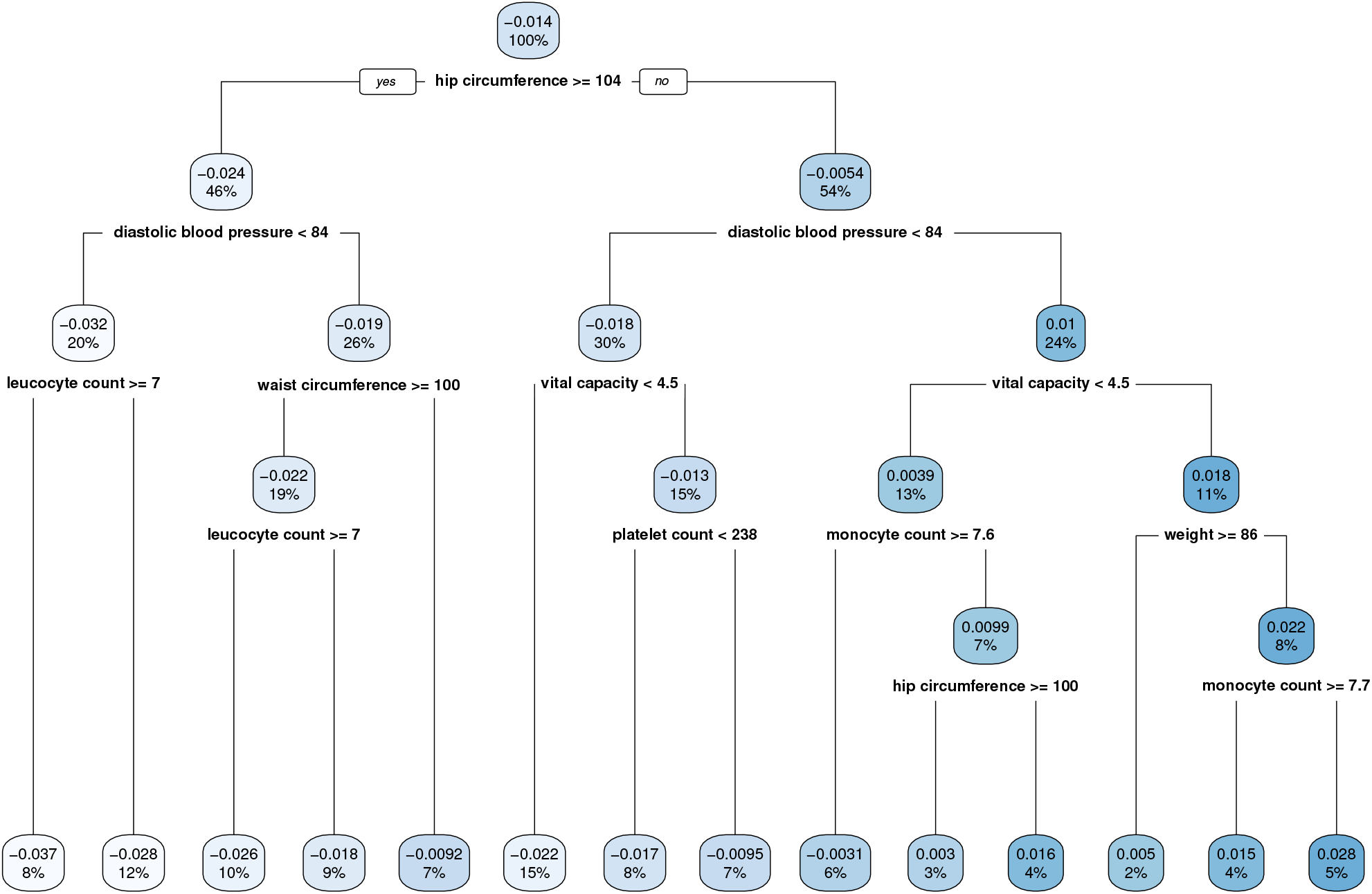
Decision tree illustrating the strata constructed by the random forest method, and the splitting criterion at each node. Values in boxes represent the average predicted treatment effect for individuals in the given node, and the proportion of the overall testing subset in that node.

## Discussion

In this paper, we have presented a non-parametric stratification method for Mendelian randomization based on a single covariate, the doubly-ranked method. We have then incorporated the stratification method into a data-adaptive approach that provides stratified estimates across a high-dimensional set of covariates. We have demonstrated the validity of our method in a simulation study, and implemented the method to show heterogeneity in the effect of BMI on lung function. We have also developed measures to assess variable importance, and to assess whether variability in individual estimates is stronger than would be expected due to chance alone.

Our applied analysis provides intriguing insights into the effect of BMI on lung function. A previous Mendelian randomization investigation demonstrated negative effects of BMI on FEV1, as well as other measures of lung function, and a positive effect on risk of asthma [40]. Another investigation found negative Mendelian randomization estimates of BMI on FEV1 that attenuated with older age [35]. We were able to show evidence that the effect of BMI on FEV1 is decreasing in BMI, but that it depends more strongly on hip circumference and weight, and less strongly on height. Taking these results at face value, this indicates that BMI has a neutral average effect on lung function in narrowly-built individuals, but a negative average effect in more broadly-built individuals. This is plausible, as the lung function of a slimmer individual may benefit from additional mass which increases physical lung capacity. However, lung function is likely to be impaired by additional mass for a plumper individual, particularly if the additional mass represents fat mass rather than muscle mass.

More generally, our method could have applications for understanding disease aetiology, particularly for the effects of complex traits that have competing effects on an outcome. There are also potential applications in terms of public health, to identify groups of the population who would benefit from an intervention, and precision medicine, to identify individuals who would benefit from a specific treatment.

The latter is most relevant to drug-target Mendelian randomization, where the exposure represents a target for pharmacological intervention [22, 13]. While ultimate arbiter of causation is the randomized trial, trials are expensive and slow to run. Our investigation can help guide trial design to focus on recruiting the most relevant population subgroups. Additionally, results of subgroup analyses in randomized trials are often controversial. Given the number of possible subgroup analyses that could be chosen, subgroup analyses can be subject to selective reporting and multiple testing [31]. Our method could be used to validate findings from subgroup analyses of randomized trials.

While the approach for stratification that we present has some novel aspects, it is a development of established techniques. Classification and regression trees, similar to the Q tree considered here, are a staple method of machine learning [24], and random forest of interaction trees have been considered previously for investigating effect heterogeneity in clinical trials [45, 46] and in Mendelian randomization [54]. Tree-based methods for causal inference, such as causal trees or causal forests, have been developed for observational studies [3, 52]. These methods typically assume the unconfoundedness condition [36], which means that all confounders are measured. In situations where confounding is a concern, instrumental variable (IV) methods can provide a natural solution. Recently, a forest for IV regression has also been discussed [4]. However, when dealing with complex scenarios where some effect modifiers may be downstream effects of the exposure, the current causal (or IV regression) tree and forest methods may not adequately address collider bias and could produce severe estimation bias, particularly if the variables used for splitting in the tree are either colliders or mediators between the exposure and the outcome [26]. The Q tree that we present is conceptually identical with other tree-based methods, but differs in its implementation as it is based on a Q statistic. The Q statistic allows a more flexible comparison of stratum-specific estimates, for example, to account for variability in the genetic effect on the exposure, as well as differential precision in stratum-specific estimates. The measures of variable importance and the permutation test that we developed in this work based on Q statistics performed well in the context of our examples.

Whereas the performance of most machine learning algorithms can be assessed directly in a testing subset, individual-level causal effects cannot be known outside of a simulation setting. This is in contrast to a typical prediction problem, where we can compare the predicted values of the outcome to its observed values. As the individual-level causal effects cannot be observed, we cannot know how well the random forest approach performs in a real-data example, as we do not know the ground truth. This means that hyperparameters, such as the minimum size of terminal node, cannot be optimally tuned to a particular applied dataset. However, in the simulation study, the variable importance measures were able to identify the key effect modifiers even without knowledge of the true effects.

There are several methodological limitations to this work. First, we create a random forest, averaging over trees that divided the population based on different covariates. While this approach will generally result in improved performance when the causal effect of the exposure depends on several covariates, it will perform less well if there is only one true effect modifier compared with a simpler approach stratifying on that covariate. Second, in calculating Mendelian randomization estimates, we make several assumptions in terms of linearity and homogeneity (or monotonicity) within strata. As Mendelian randomization estimates represent the impact of a lifelong shift in the distribution of an exposure [20], we generally do not encourage an overly literal interpretation of Mendelian randomization estimates as causal effects that are achievable in practice [12]. Additionally, Mendelian randomization estimates may depend on the specification of the Q tree, the causal interpretation of the Mendelian randomization estimate depends on the choice of stratifying covariates. We assume that the relative magnitude of different Mendelian randomization estimates in subgroups is indicative of the relative magnitude of the effect of intervention on the exposure in the same subgroups in practice. Third, all the covariates that we have considered are continuous variables. While the doubly-ranked method can be used for a discrete covariate, caution should be taken, particularly when dividing into a large number of strata if the covariate takes a small number of values. Fourthly, the doubly-ranked method makes the rank-preserving assumption, which cannot be tested empirically. Finally, as with all Mendelian randomization investigations, results are dependent on the validity of the genetic variants as instrumental variables.

There are also limitations to the applied analysis. As with most cohorts, UK Biobank is known to suffer from selection bias, the magnitude of which depends on the age of participants [21]. UK Biobank participants of working age are more likely to be affluent, early retirees, and so the stratification on age at recruitment is likely to reflect differences in socio-economic status in addition to age. Some individuals were not able to blow into the spirometer, or were not able to provide a reliable measure, and so have been excluded from the analysis; this could also lead to selection bias. In order to avoid variability in estimates due to sex-based differences in the distribution of BMI and other anthropometric traits, we restricted analyses to include men only. Additionally, to avoid population stratification, we restricted analyses to individuals of European ancestries. This means our results may not be generalizable to other population groups.

In summary, our data-adaptive method can investigate effect heterogeneity in the effect of an exposure on an outcome in a Mendelian randomization framework. This can provide important insights into disease aetiology, and into finding groups of individuals who would most benefit from intervention on the exposure.

## Methods

### Modelling assumptions and estimands

Our focus lies on effect estimation conditional on covariate information. These effects are also referred to as conditional average treatment effects (CATE):

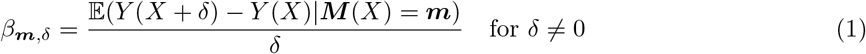

where *Y* (*x*) represents the potential outcome with the exposure level *x* (following the potential outcome framework [33, 37]), *X* is the continuous exposure (for binary exposure, we define *β*_***m***_ = 𝔼(*Y* (1) *− Y* (0)|***M*** (*X*) = ***m***) accordingly), and ***M*** is the high-dimensional covariate [3, 52]. The average causal effect could be modified by the covariate level ***m***, and hence the CATE given ***M*** (*X*) = ***m*** is possibly heterogeneous. A non-linear causal effect is a special example of heterogeneous effect where the exposure level itself acts as an effect modifier.

The general model is expressed as a DAG in the left panel of Table 1. It is required that *Z* is a valid instrument, which means that when *M → Y* exists, there is no direct causal path from *Z* to *M*. Additionally, a path *Z → M* indicates that *M* is a collider, regardless of the specific relationship between *X* and *M*. If the covariate *M* is either a collider or a mediator, stratifying naively on *M* will violate the exchangeability assumption, resulting in a biased CATE estimator.

**Table 1:**
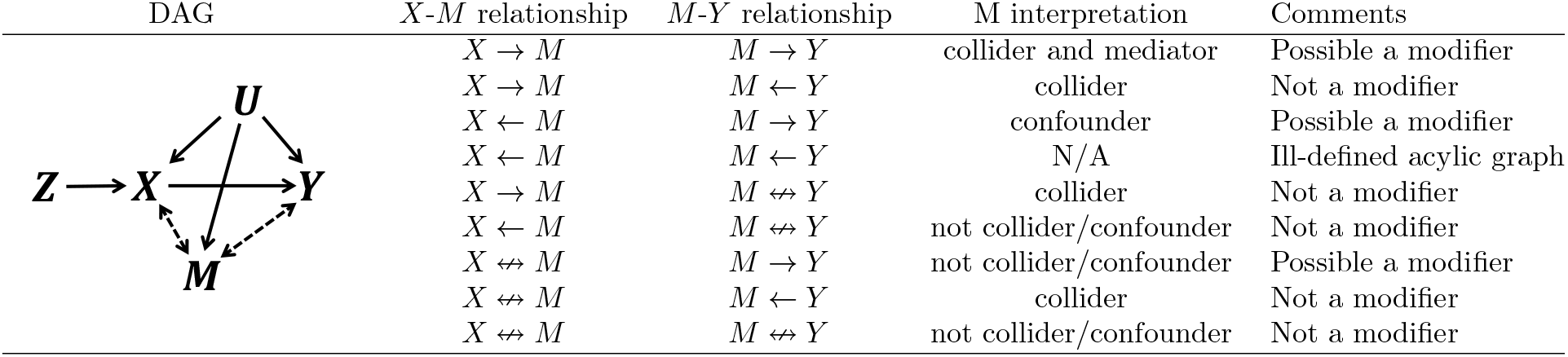
Possible effect heterogeneity scenarios. The left panel is a directed acyclic graph (DAG) where *Z,X,Y,U* represent the instrument, the exposure, the outcome, and the unmeasured confounders, respectively. *M* is a variable to be considered. The DAG has different possible scenarios, each of which has different arrow directions (or no arrow, denoted by ↮) for the *X-M* and *M-Y* relationship.

It is important to note that the observed covariates, ***M***, are not necessarily the true effect modifiers that demonstrate the heterogeneity of CATE. This is because some covariates may be correlated with unmeasured true effect modifiers. However, the observed covariates are still valuable if the objective is to predict the individual effect given its covariate information.

### Collider robust stratification

Collider bias is widespread in the analysis of observational data and is challenging to causal inference [34, 32]. Some typical sources of collider bias include: (i) selection bias occurring when selection into a study sample depends on a collider [25]; (ii) survivor bias occurring when survival depends on a collider [6, 42]; and (iii) in an instrumental variable analysis, conditioning on the exposure directly [28], as the exposure is a function of the instrument and confounders, and hence a collider. Collider bias can also occur in an instrumental variable analysis when stratifying on a covariate, if the covariate is a function of the instrument and confounders. As the exposure is a function of the instrument and confounders, any covariate causally downstream of the exposure will be a collider. Even if the instrumental variable assumptions are satisfied for the population as a whole, they are typically invalid within strata of the population defined by a collider [14, 23].

The residual stratification method derives the counterfactual value of a covariate *M* in a parametric model. The method assumes that the structural equation for the covariate is linear and homogeneous in the instrument *Z*: *M* = *M* (0) + *αZ*, where the counterfactual variable *M* (0) can be estimated by taking the residuals from regression of *M* on *Z*. We then form strata based on these residual values *M* (0). As *M* (0) is not a function of the instrument *Z*, it is typically not a collider even if *M* is a collider [17].

The doubly-ranked method is a nonparametric stratification method that relaxes the assumptions of the residual method. The method has previously been described for stratifying on the exposure, in an approach known as non-linear Mendelian randomization [50]; we here adapt the method to stratify on a covariate. We assume that the sample size is *N* = 10 *× K*, and the number of strata desired is two. The method is performed by the following steps:

1. Rank individuals according to their value of the instrument, and form *K* pre-strata of size 10 by stratifying on the instrument. Ties are broken at random.
2. Rank individuals within each pre-stratum based on their value of the covariate.
3. Form the two strata by selecting the individuals with the same covariate rank range from each pre-stratum; such that stratum 1 contains the individuals with the lower 5 values of the covariate from each pre-stratum and stratum 2 contains the individuals with the higher 5 covariate values from each pre-stratum.

This method stratifies the population using information on a covariate under a rank preserving assumption. We assume that each individual’s counterfactual values of the covariate have the same rank ordering for different values of the instrument. This assumption is illustrated for a dichotomous instrument (*Z* = 0, 1) in Figure 7. The black line illustrates the distribution of the covariate for those with *Z* = 0, and the blue line illustrates the distribution of the covariate for those with *Z* = 1. For instance, we consider an individual with *Z* = 0 and covariate value equal to the 10th percentile of the covariate distribution for those with *Z* = 0. If this individual instead had *Z* = 1, we assume that their value of the covariate would be at the 10th percentile of the covariate distribution for those with *Z* = 1. The linear and homogeneous model required by the residual method is a special case of this assumption. We refer to an individual’s quantile in the relevant covariate distribution as their rank index. The rank index defines the potential values of the covariate at different values of the instrument (all but one of which will be counterfactual).

**Figure 7:**
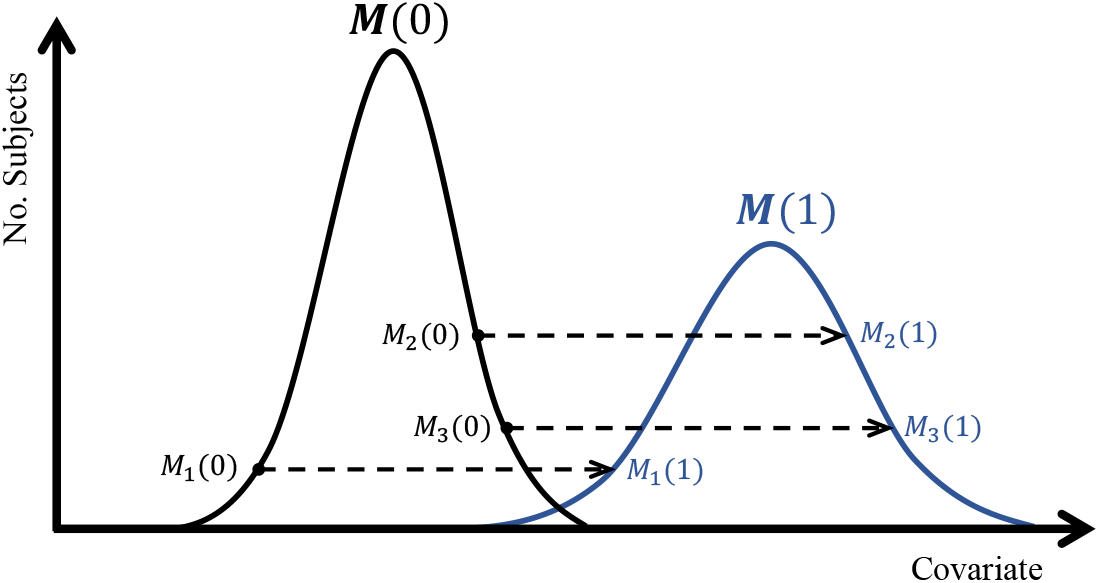
Diagram illustrating the rank preserving assumption for a dichotomous instrumental variable *Z* with counterfactual covariate distributions ***M*** (0) (the black group) and ***M*** (1) (the blue group). The dashed arrow represents the one-to-one mapping from the counterfactual covariate value with *Z* = 0 to the counterfactual covariate value with *Z* = 1.

The first step of the doubly-ranked method divides the population into pre-strata, such that individuals in the same pre-stratum have similar values of the instrument, and so ordering by the covariate within the pre-stratum approximates the rank index of individuals. By selecting individuals according to their rank in the pre-strata, we obtain strata with different average levels of the covariate, but a wide range of values of the instrument. As the rank index is not a function of the instrument, this stratification will not induce collider bias.

### Assessing heterogeneity in stratum-specific estimates

Having constructed strata using the residual or doubly-ranked method, we evaluate a measure of heterogeneity across the stratum-specific Mendelian randomization estimates. We calculate the association of the genetic instrument with the exposure in stratum *k* as 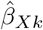 with standard error *σ*_*Xk*_, and the association of the genetic instrument with the outcome in stratum *k* as 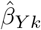 with standard error *σ*_*Y k*_. The stratum-specific causal estimates are obtained using the ratio method as 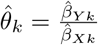. Cochran’s Q statistic can be obtained as [15]

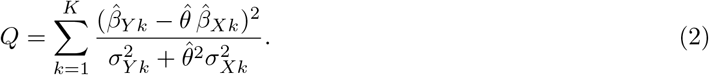

where 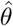 is the inverse-variance weighted average of the stratum-specific estimates. Under the null hypothesis that the stratum-specific estimates are all targeting the same parameter, the Q statistic should have a 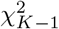 distribution, where *K* is the number of strata. A higher Q value gives stronger evidence of heterogeneity between stratum-specific estimates, therefore indicating greater effect modification by the covariate used for stratification. Note that the Q statistic can be understood as a component of the profile log-likelihood considering the uncertainty of the estimated instrument-exposure association and is robust to weak instruments [7, 56], which is important as weak instruments may be common within strata, due to the reduced sample size.

### Building a single Q tree

As the covariate information for many applications is high-dimensional, stratification on all covariates may be infeasible, and data-adaptive methods may be preferable to stratification on a small number of selected covariates. As a simple but powerful method, the Q tree method can help to build strata considering multiple covariates in an agnostic way. When utilizing tree or random forest methods, it is crucial to define the rules for recursive partitioning that can effectively detect and emphasize the heterogeneity related to the specific aspect of interest in the study [4]. In this context, we leverage the collider-robust partitioning approach to avoid collider bias in IV analysis. Additionally, we incorporate the Q heterogeneity statistics, which have been widely employed to assess heterogeneity in MR studies. Starting with the initial node containing all participants to be stratified, a Q tree is constructed by the following steps:

1. **Determine the splitting covariate**. We form two strata for the present node based on each candidate covariate using a stratification method. The splitting covariate *M* for the present node and the splitting proportion are chosen to give the greatest Q statistic value.
2. **Split based on the candidate covariate**. Two child nodes are built based on the selected splitting covariate *M* and splitting proportion. We then either return to step 1 to split each child node, or stop if the stopping rule is met.

The stopping rule is either: (1) the greatest Q statistic value is less than 3.84 (the 95th percentile of a chi-squared statistic with one degree of freedom); (2) the size of the child node is less than 1,000; or (3) the single node depth is larger than 5. We consider three possible splitting proportions, expressed as the ratio of the sub-node sizes: namely, 3 : 7, 5 : 5, and 7 : 3. The end nodes are the strata for Mendelian randomization analysis. Note that the same splitting covariate may be selected multiple times. Such an algorithm is quite similar to the simple yet powerful tree method CART (Classification And Regression Tree), but the splitting and stopping rules in our algorithm are based on Q statistic; therefore we call it a Q tree.

Once we have built a Q tree, it can be used to predict causal effects for individuals in the testing subset. This is completed by passing this individual down the fitted Q tree. At each branch, the individual will go to the sub-strata of which the mean values of *M*^*\**^ is closer to this individual’s *M*^*\**^ value, where *M*^*\**^ is the chosen splitting covariate for that node. Specifically, the decision rule can compare the value of *M*^*\**^ to the boundary value

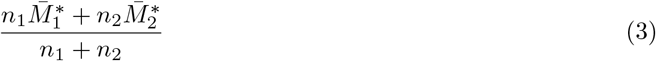

where *n*_1_ and *n*_2_ are the sample size of the lower and upper sub-node, respectively; 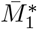 and 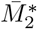 are the corresponding mean values of the chosen covariate in the sub-nodes. Once this individual reaches the end node, the predicted treatment value is the stratum-specific Mendelian randomization estimate for that stratum.

An important concept in tree-based estimation is honesty. In honest estimation, separate subsets of the training data are used to construct the tree and evaluate the node estimates [3, 26]. Honesty leads to favorable properties in terms of convergence and inference [3, 52]. However, honest estimation may not be suitable for our Q tree. This is because when constructing sub-groups within the estimation data based on a tree, decision rules typically involve covariate stratification, which can introduce collider bias and lead to biased leaf-specific estimators for the estimation data. Therefore, we use the subgroups generated by the doubly-ranked stratification, where the IV assumptions hold and collider bias is avoided, to obtain the leaf-specific estimates. For this reason, we only consider a limited number of splitting proportions for each covariate, to avoid overfitting within the training subset.

### Building a random forest of Q trees

The random forest is a bootstrap aggregating (bagging) method for reducing an estimator’s variance by aggregating multiple de-correlated trees [8, 24]. To construct a random forest of Q trees (RFQT), we first take *N*_*B*_ bootstrap samples of the training subset, where *N*_*B*_ is the size of the forest. For each bootstrapped dataset, we build a Q tree as introduced before, but at each split, only a random set of covariates are considered as candidate covariates for that node. In the simulation study and applied example, we consider 40% of covariates at each division. The RFQT estimate for any individual is the average predicted value from all the Q trees. The final forest size *N*_*B*_ is chosen such that the OOB error and the test error (if applicable) of the RFQT are stable as the number of trees increases (see Supplementary Figure S5). Unlike most supervised learning problems, the real data in our context do not have relevant labels (i.e. individual-level causal effects), which means the tuning parameters such as *N*_*B*_ and the proportion of covariates considered at each division cannot be directly determined by the OOB or testing subset error. The forest size *N*_*B*_ for the real data fitting is therefore chosen such that individual predicted effects are converged.

### Variable importance

Variable importance (VI) measures which covariates contribute to the predictive accuracy of effect estimates. VI is an example of a cost-of-exclusion approach and can be well-compatible with tree and random forest models [27]. The VI measurement in a RFQT is obtained by OOB samples from each Q tree boot-strap by the algorithm 1, which is similar to that previously proposed for an interaction tree [45]. In a simulation study, the prediction accuracy can be the MSE of the individual-level effect estimates. The change of accuracy can be the difference between the MSE before and after the permutation. With real data, the change of accuracy is replaced by the change in effect estimates for the OOB samples after permuting the covariate, compared with using the unpermuted sample. That is, 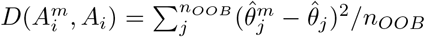 for the *m*-th covariate where 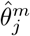 and 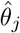 are the *j*-th individual effect estimates by using the Q tree 𝒬_*i*_ with the OOB samples 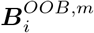 and 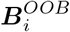, respectively; and *n*_*OOB*_ is the number of OOB samples. More important variables in both the simulation and real application should contribute to a greater change of accuracy.

#### Algorithm 1 Computing Variable Importance (VI) Measure of RFQT

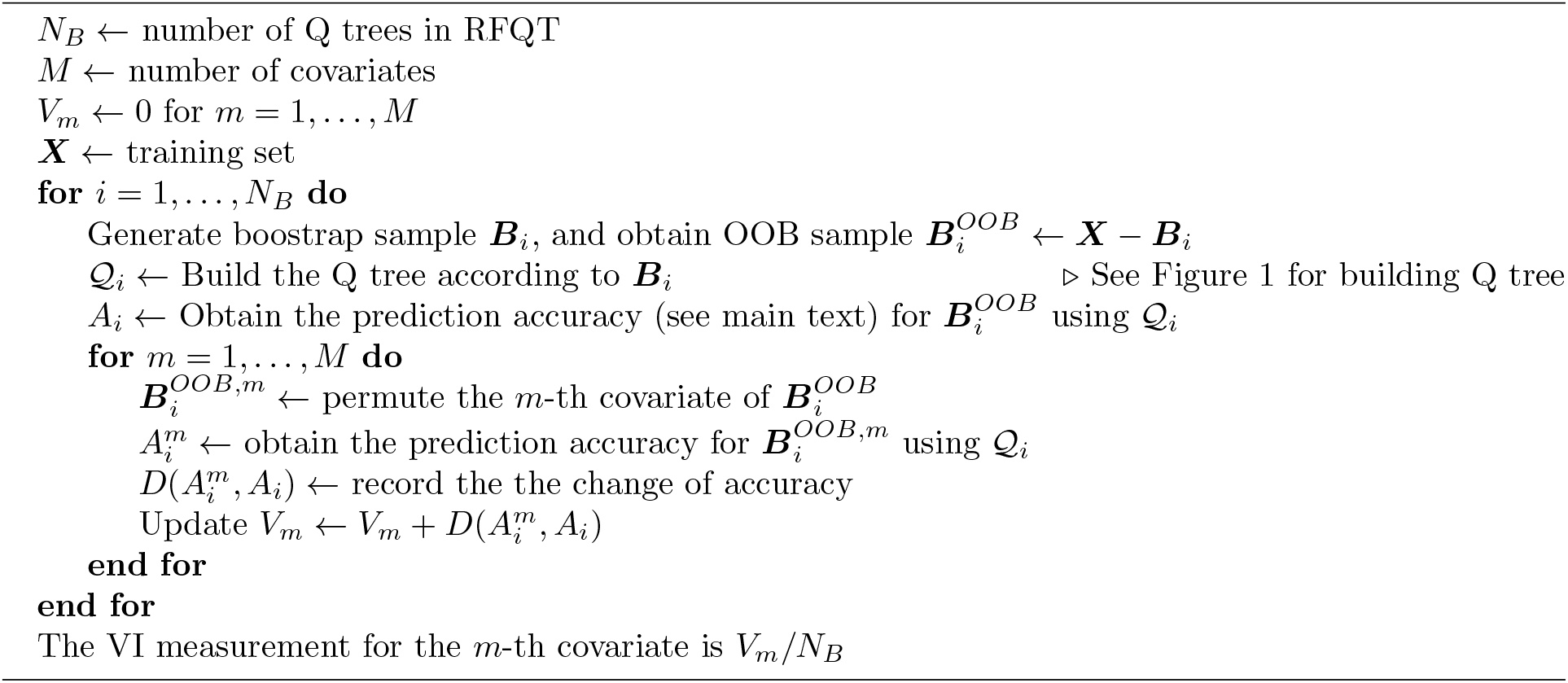

### Permutation test of heterogeneity

We propose two nonparametric permutation test statistics for assessing effect heterogeneity of predicted estimates in the training dataset, accounting for uncertainty from the RFQT algorithm. The null hypothesis is that all the candidate covariates considered do not modify the treatment effect, and so the individual-level predicted causal effects are not more variable than would be expected due to chance alone.

We randomly permute the candidate covariates in the training subset to mimic the null scenario. For each permutation, we build the RFQT for the permuted sample and derive the test statistics *S*_1_, which is similar to that proposed in a previous paper [54],

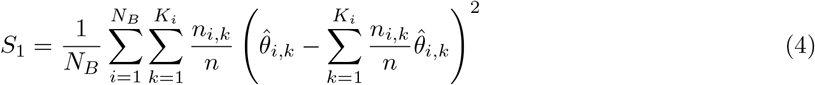

and *S*_2_, which considers variability in the instrument–exposure association estimates:

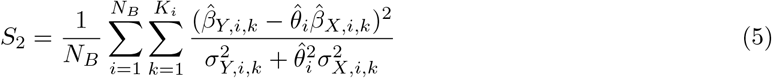

where *N*_*B*_ is the number of Q trees, *n* is the training sample size, *n*_*i,k*_ is the size of the *k*-th end node (strata) in the *i*-th Q tree that satisfies 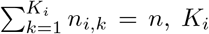 is the number of end strata for the *i*-th Q tree, 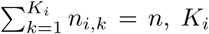 is the MR estimate for the *k*-th end strata in the *i*-th Q tree, 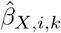 and 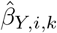 are the estimated instrument-exposure and instrument-outcome association respectively for the *k*-th strata of the *i*-th Q tree. *σ*_*X,i,k*_ and *σ*_*Y,i,k*_ are their corresponding standard errors. 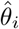 is the inverse-variance weighted average of the stratum-specific estimates for the *i*-th Q tree. The larger value of *S* gives stronger evidence to reject the null hypothesis. Compared with *S*_1_, *S*_2_ is more robust to extreme values caused by weak instruments as it allows for the variability in the instrument–exposure association estimates.

For each permutation test statistic, we derive the an empirical p-value, which is the proportion of permuted datasets having a larger value of the test statistic than that from the original data, to decide if the candidate covariates as a whole modify the treatment effect.

### Reducing variability in stratum-specific estimates

One notable characteristic of the doubly-ranked method is that the stratification results can exhibit high variability due to the ranking process. This high variability can be mitigated by employing resampling procedures, such as random forest, which can help stabilize the results. However, when using a one-time doubly-ranked stratification approach, this variability may impact the results. To address this issue and obtain more stable stratum-specific estimates, we utilize a resampling procedure similar to Rubin’s rules [38, 30] to reduce the variability and improves the stability of the estimated stratum-specific effects.

Given a dataset for fitting, we employ a multiple sampling approach where we randomly exclude 10 individuals in each iteration. This random omission sufficiently alters the individual rank information, consequently affecting the stratification results. Let’s denote the total number of sampling times as *S*. In each sampling iteration, we obtain the stratum-specific estimates: 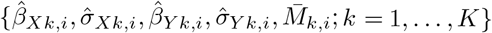 for the sampling time *i* = 1, 2, …, *S*, where 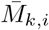 represents the average covariate value for the *k*-th stratum in the *i*-th sampling time. We obtain the pooled point estimator for the *k*-th stratum instrument-outcome association

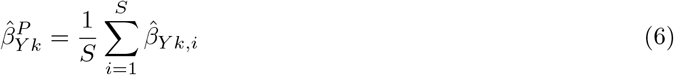

and its 95% confidence interval, (*L*_*Y k*_, *R*_*Y k*_):

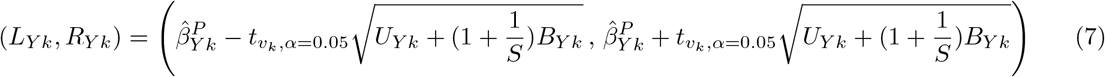

where 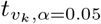 represents the 0.975 quantile point of the t distribution with *v*_*k*_ degrees of freedom, 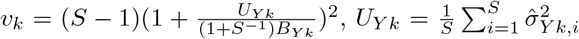, and 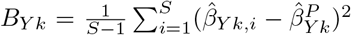. We can also obtain similar pooled estimates for the instrument-exposure associations. Therefore, we have the stratum-specific pooled MR estimates as 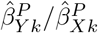 with the 95% confidence interval 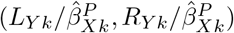. The corresponding covariate value on the x-axis is 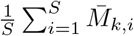. We therefore calculate the Q statistic with the pooled estimates as

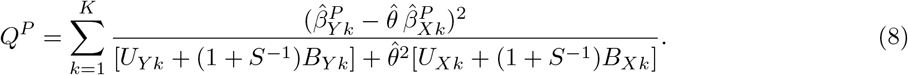

where 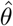 is the inverse-variance weighted average of the stratum-specific pooled estimates. We also test the trend association of the pooled stratum-specific estimates against the stratum-specific covariate values via meta-analytic mixed-effects models, where the stratum-specific covariate values are considered as the moderator explaining the heterogeneity [5]. The test can be implemented using the R package *metafor* [51].

### Simulation study

To compare the performance of the stratification methods, as well as the RFQT method, we conduct a simulation study considering the following data-generating model, where the individual index has been omitted for notational brevity,

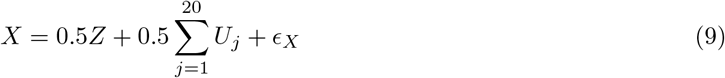

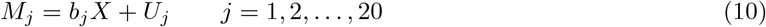

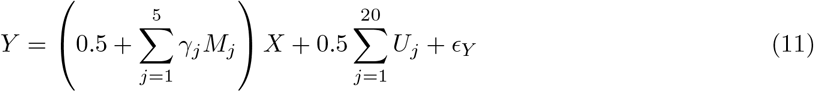

where *Z, X, {U*_*j*_*}, {M*_*j*_*}* and *Y* are the instrument, the exposure, unmeasured confounders, the candidate covariates, and the outcome, respectively. 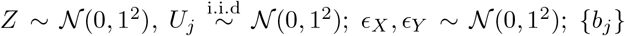 are the effects of the exposure on each candidate covariate; *{γ*_*j*_*}* are the modifier effects by each candidate covariate and 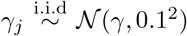 otherwise. That is, the first five covariates are effect modifiers. We call *γ* the strength of modification. Note that even for *γ* = 0, there is weak effect modification.

We consider three scenarios for the effects of the exposure on the candidate covariates

A: *b*_*j*_ = 0, *j* = 1, 2, 3, …, 20

B: *b*_*j*_ = 0.5 when *j* = 2, 4, 6, …, 20 and 0 otherwise

C: *b*_*j*_ = 0.1 + 0.5*U*_*j*_, *j* = 1, 2, …, 20

In Scenario A, all the candidate covariates are not colliders. In Scenario B, half of the covariates are colliders, as they are common effects of the exposure and the unmeasured confounders. Scenario C corresponds to a more complex case in which the effects on the candidate covariates are modified by the unmeasured confounders, so both of the the assumptions requied by the residual method and the rank preserving assumption are violated. The simulation scenarios can be expressed by the DAG in Supplementary Figure S6.

When calculating the mean squared error of predicted estimates, we compare the individual-level predicted estimates to the controlled direct effects of the exposure for an individual’s values of the covariates: this is 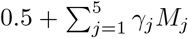. This is for computational reasons: it is simpler to calculate than the total effect of the exposure (which is the quantity targeted by the Mendelian randomization estimates), but the quantities should be close in practice.

### Applied example: body mass index on lung function

In order to implement RFQT in a real application, we took data on 167,121 male individuals from UK Biobank (Supplementary Figure S7). A weighted gene score comprising 94 uncorrelated (pairwise *r*^2^ *<* 0.01) single nucleotide polymorphisms (SNPs) was used as an instrumental variable. These SNPs have previously been shown to be associated with BMI at a genome-wide level of statistical significance [29]. This genome-wide association study did not include UK Biobank participants, thus avoiding bias due to winner’s curse [48]. Weights for the gene score were obtained from UK Biobank participants. We took BMI as the exposure of interest and FEV1 as the outcome of interest. We used 27 other distinct variables and the exposure itself (to consider a potential non-linear pattern) as candidate covariates.

We took two-thirds of individuals as the training subset and the remaining one-third of individuals as the testing subset. For the RFQT method, we chose the number of trees to be 200, as it was found that 100-200 trees was sufficient for converged predicted values. We use similar hyperparameters for RFQT as in the simulation study; that is, an end node size of 1,000, a maximum tree depth 5, and a threshold Q value of 3.84 for the stopping rules. For each node in each tree, a random subset of 11 variables (i.e. around 40%) were considered as candidate splitting covariates. We used the doubly-ranked stratification method. Variable importance measurements were recorded for all the covariates. We applied the permutation test for the permutation test statistics *S*_1_ and *S*_2_ by permuting covariate information for the training subset 1000 times. The empirical p-values were 0.058 (95% CI: 0.045, 0.074) for *S*_1_ and *<* 0.001 (95% CI: 0.000, 0.003) for *S*_2_. The confidence intervals are derived by the logistic regression model and the rule of three, respectively. The test result suggests that *S*_2_ was a more discriminating measure of effect heterogeneity in this example.

## Data Availability

The individual-level data underlying the applied example presented in the study is not publicly available due to privacy policies. However, it can be accessed by researchers through an application to UK Biobank (https://www.ukbiobank.ac.uk). The fitted data used for generating the results in both the simulation and applied example are provided at https://github.com/HDTian/RFQT/tree/main/Data

## Availability of data and materials

R-code for the simulation study and real application (R version ⩾ 4.2.2, MIT license) is available at https://github.com/HDTian/RFQT. The illustration codes are provided on https://github.com/HDTian/RFQT/tree/main/illustration_sim_real, which allows the reader to reproduce all results and adapt the presented methodology to their own research.

## Acknowledgements

This work was supported by the Wellcome Trust (225790/Z/22/Z) and the United Kingdom Research and Innovation Medical Research Council (MC UU 00002/7). B.D.M.T. is supported through the United Kingdom Medical Research Council programme grant MC UU 00002/2. S.B. is supported by a Sir Henry Dale Fellowship jointly funded by the Wellcome Trust and the Royal Society (204623/Z/16/Z). The funders had no role in study design, data collection and analysis, decision to publish, or preparation of the manuscript. The research has been conducted using the UK Biobank Resource under Application Number 98032. For the purpose of open access, the author has applied a Creative Commons Attribution (CC BY) licence to any Author Accepted Manuscript version arising from this submission.

## Author contributions

H.T. and S.B. conceived and designed the study. H.T carried out the statistical and computational analyses. B.D.M.T. and S.B. contributed to the design of the study and the interpretation of the findings. The paper was written by H.T. and S.B.; and revised by all the co-authors. All co-authors have approved of the final version of the paper.

## Supplementary Materials

### Supplementary Table

**Supplementary Table S1:** Abbreviations of the variables considered in the UK Biobank application example. As body mass index is the exposure, stratifying on body mass index is equivalent to a non-linear Mendelian randomization analysis.

| Abbreviation | Meaning |
| --- | --- |
| hip | Hip circumference |
| wt | Weight |
| dbp | Diastolic blood pressure |
| waist | Waist circumference |
| monos | Monocyte count |
| leuc | Leucocyte count |
| vitcap | Vital capacity |
| bmi.1 | Body mass index |
| pef | Peak expiratory flow |
| whtr | Waist to height ratio |
| sbp | Systolic blood pressure |
| neutros | Neutrocyte count |
| urianac | Urinary sodium |
| whr | Waist to hip ratio |
| uriamac | Urinary microalbumin |
| handgpr | Hand grip strength – right |
| eosins | Eosinophil count |
| hemcrit | Haematocrit |
| lymphs | Lymphocyte count |
| uriakc | Urinary potassium |
| pulse | Pulse rate |
| ages | Age at survey |
| handgpl | Hand grip strength – left |
| uriacc | Urinary creatinine |
| ht | Height |
| platelet | Platelet count |
| rbc | Red blood cell count |
| hemglob | Haemoglobin |

### Supplementary Figure

**Supplementary Figure S1:**
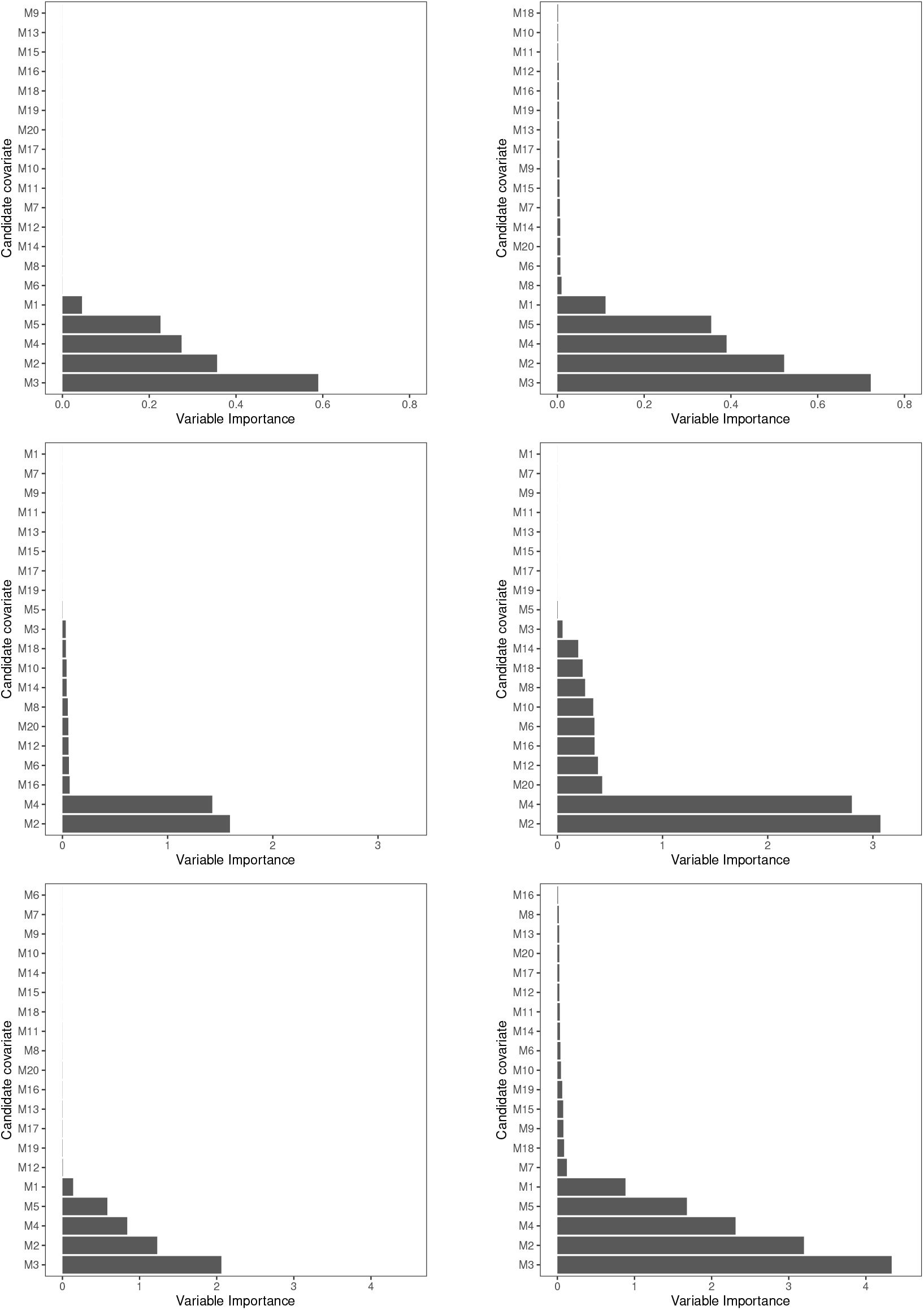
Variable importance (VI) measurements for the doubly-ranked method with random forest. Left panels: VI measurements calculated using individual effect labels (that is, the true individual effects), based on changes in mean squared error. Right panel: VI measurements calculated without individual effect labels, based on changes in estimates. The top, middle, and bottom results correspond to a single randomly chosen simulated dataset under scenario A, B, and C with the strength of modification 0.5, respectively. The true effect modifiers are M1-M5.

**Supplementary Figure S2:**
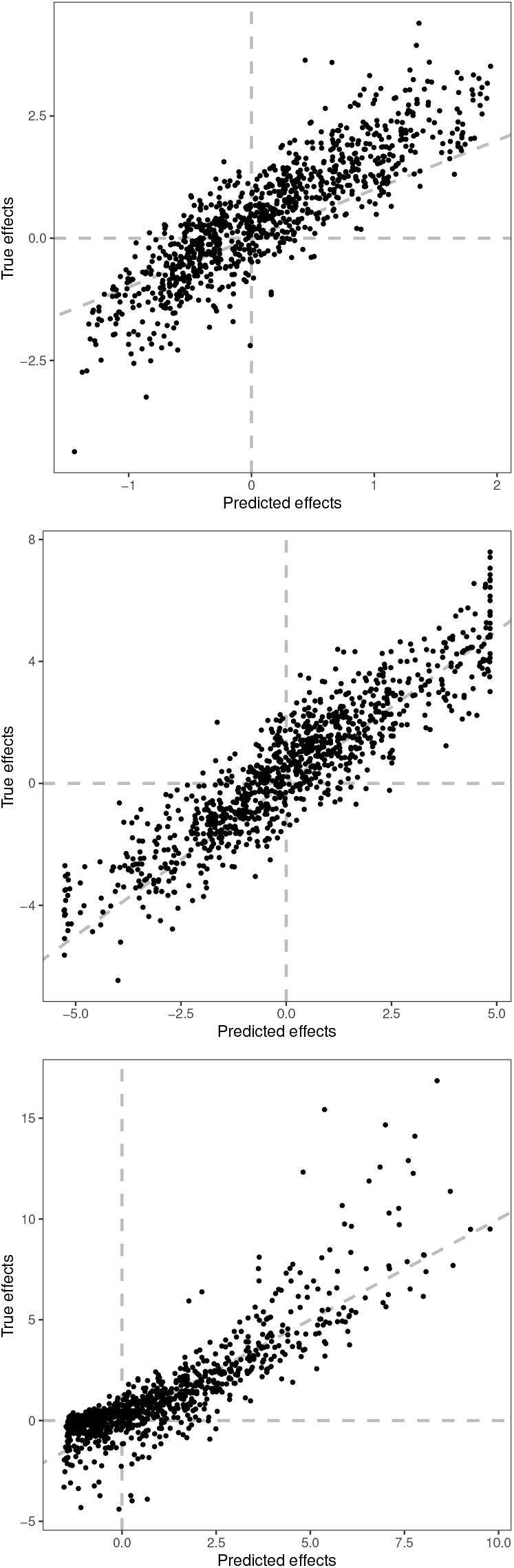
Scatterplot displaying the predicted effects and true effect values for 1000 randomly selected individuals from the testing set for the doubly-ranked method with random forest. The top, middle, and bottom plots correspond to a single randomly chosen simulated dataset under the simulation scenarios A, B, and C with a strength of modification of 0.5, respectively.

**Supplementary Figure S3:**
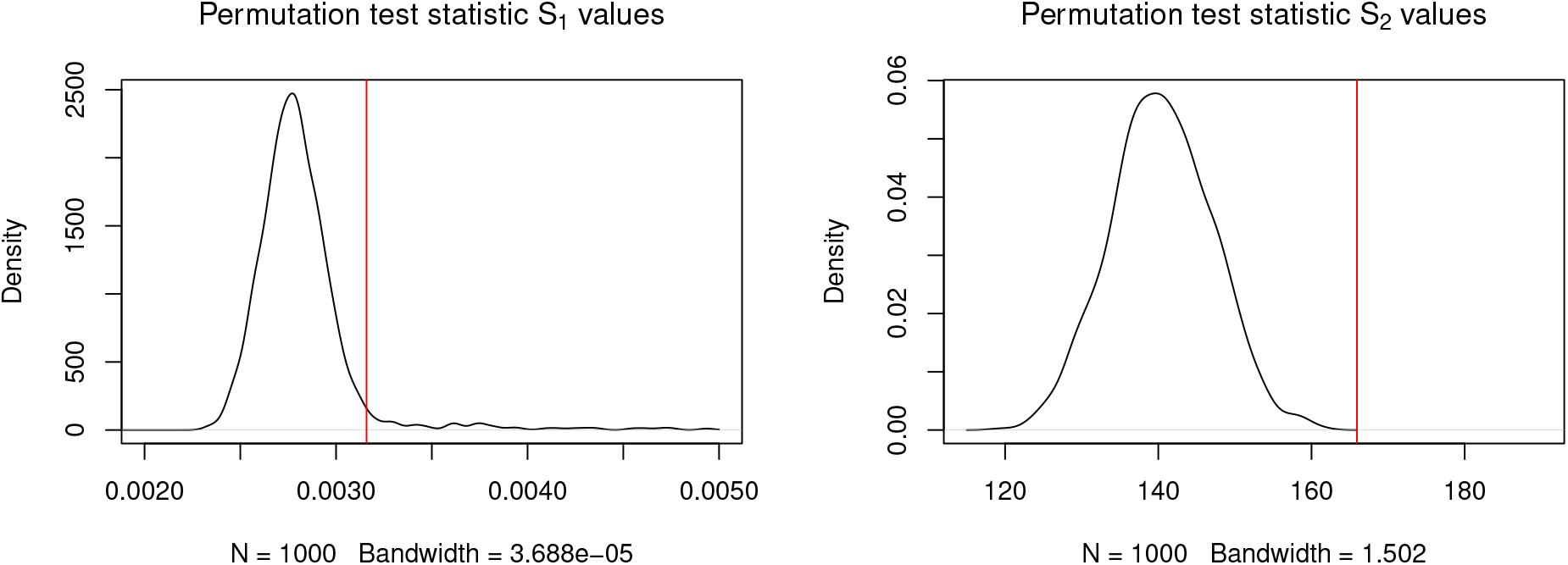
The kernel smoothed density of the under-null samples of the permutation test statistics *S*_1_ (left) and *S*_2_ (right) with 1000 permutations. The statistic values for the unpermuted data are shown by the red lines. The bandwidth is decided by Silverman’s rule of thumb.

**Supplementary Figure S4:**
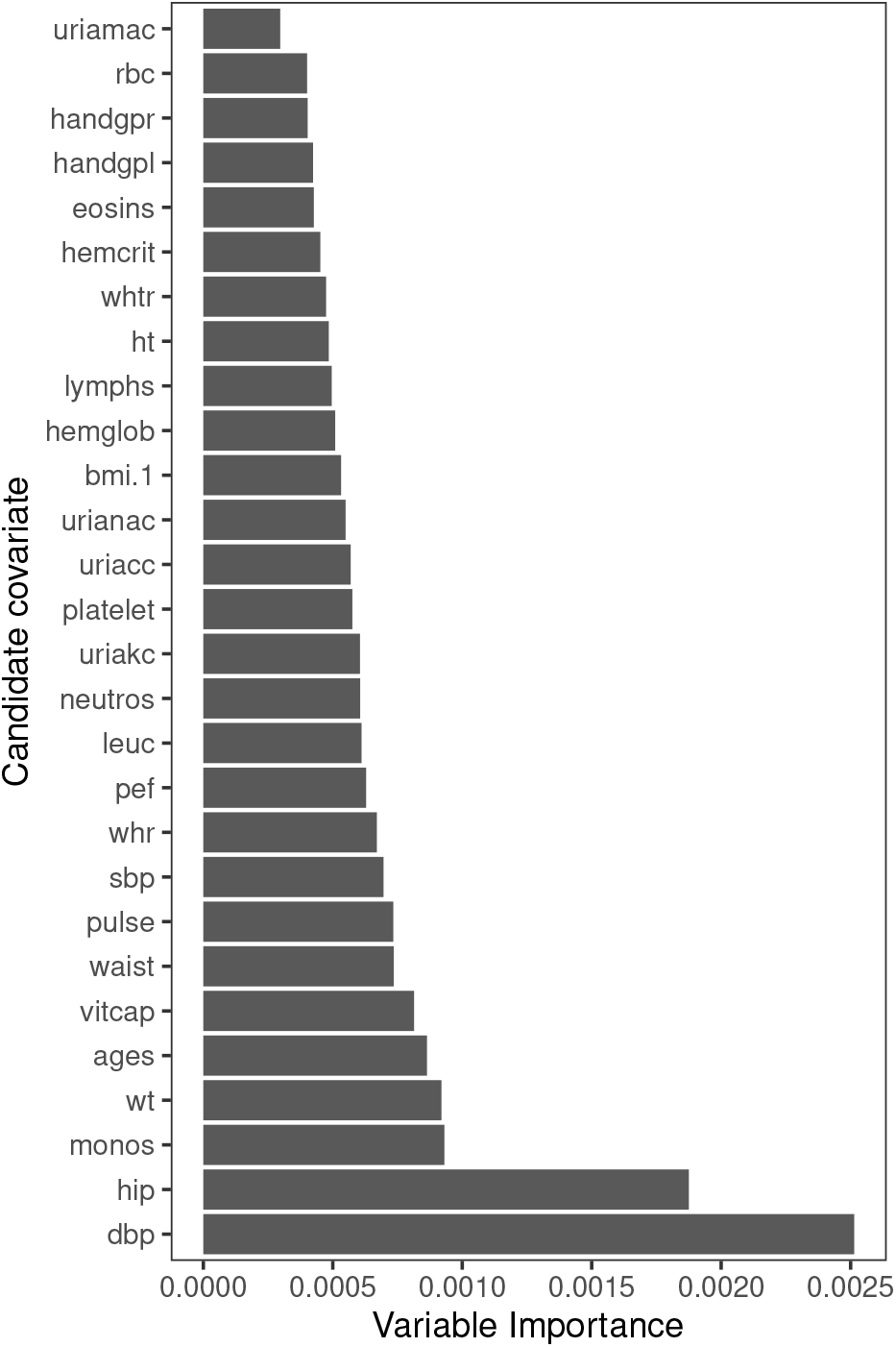
Variable importance measures in the UK Biobank example for 28 candidate covariates.

**Supplementary Figure S5:**
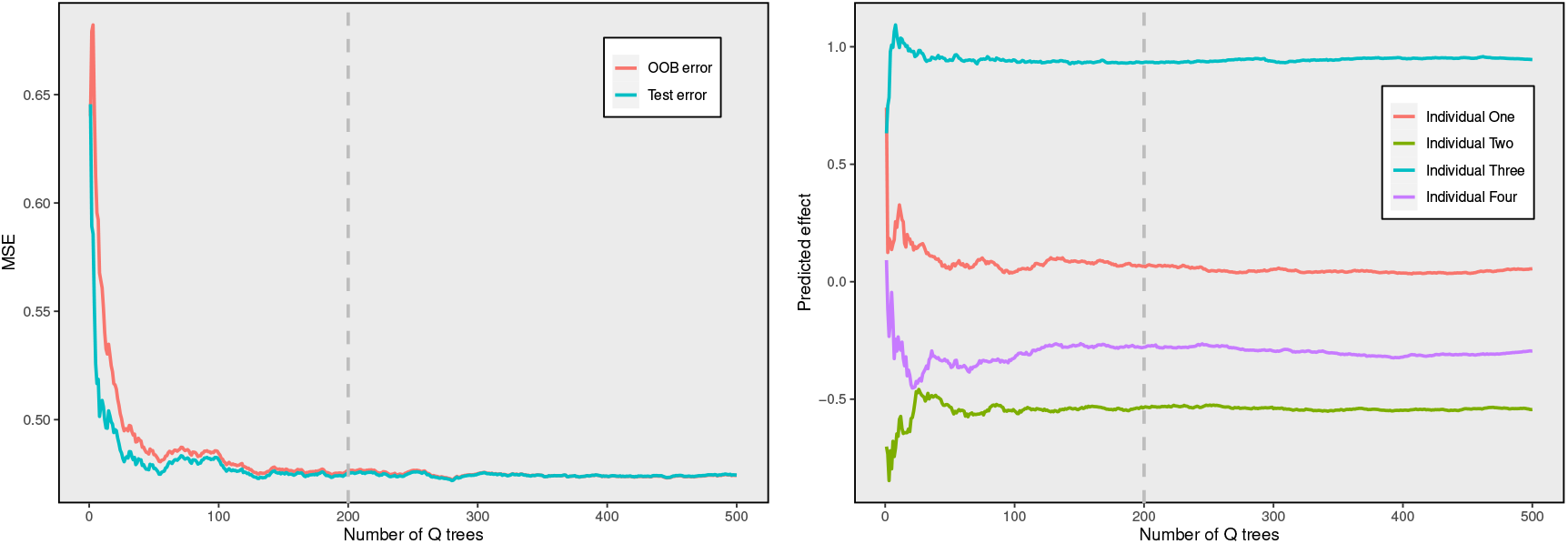
Left panel: the mean squared error (MSE) of the OOB samples (red curve) and testing subset samples (blue curve) with increasing numbers of Q trees for the simulation study. Right panel: the predicted values of four randomly selected samples of the testing subset with increasing numbers of Q trees for the simulation study. OOB: out-of-bag.

**Supplementary Figure S6:**
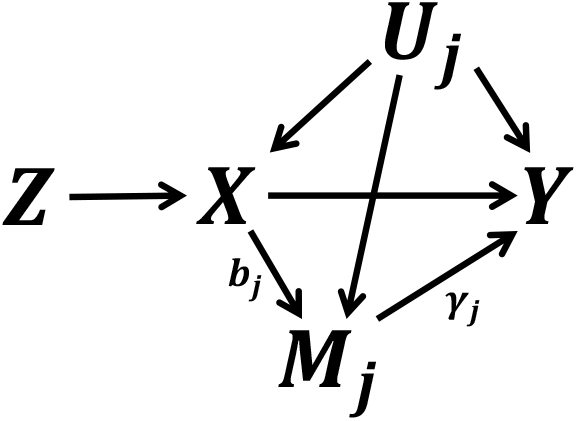
Directed acyclic graph (DAG) demonstrating the variable relationships in simulation. *Z, X, Y, U*_*j*_, *M*_*j*_ represents the instrument, the exposure, the outcome, the *j*-th confounders and the *j*-th covariate. *b*_*j*_ represents the effect of the exposure on the *j*-th covariate, and *γ*_*j*_ represents the modification effect of the *j*-th covariate on the direct effect of the exposure on the outcome.

**Supplementary Figure S7:**
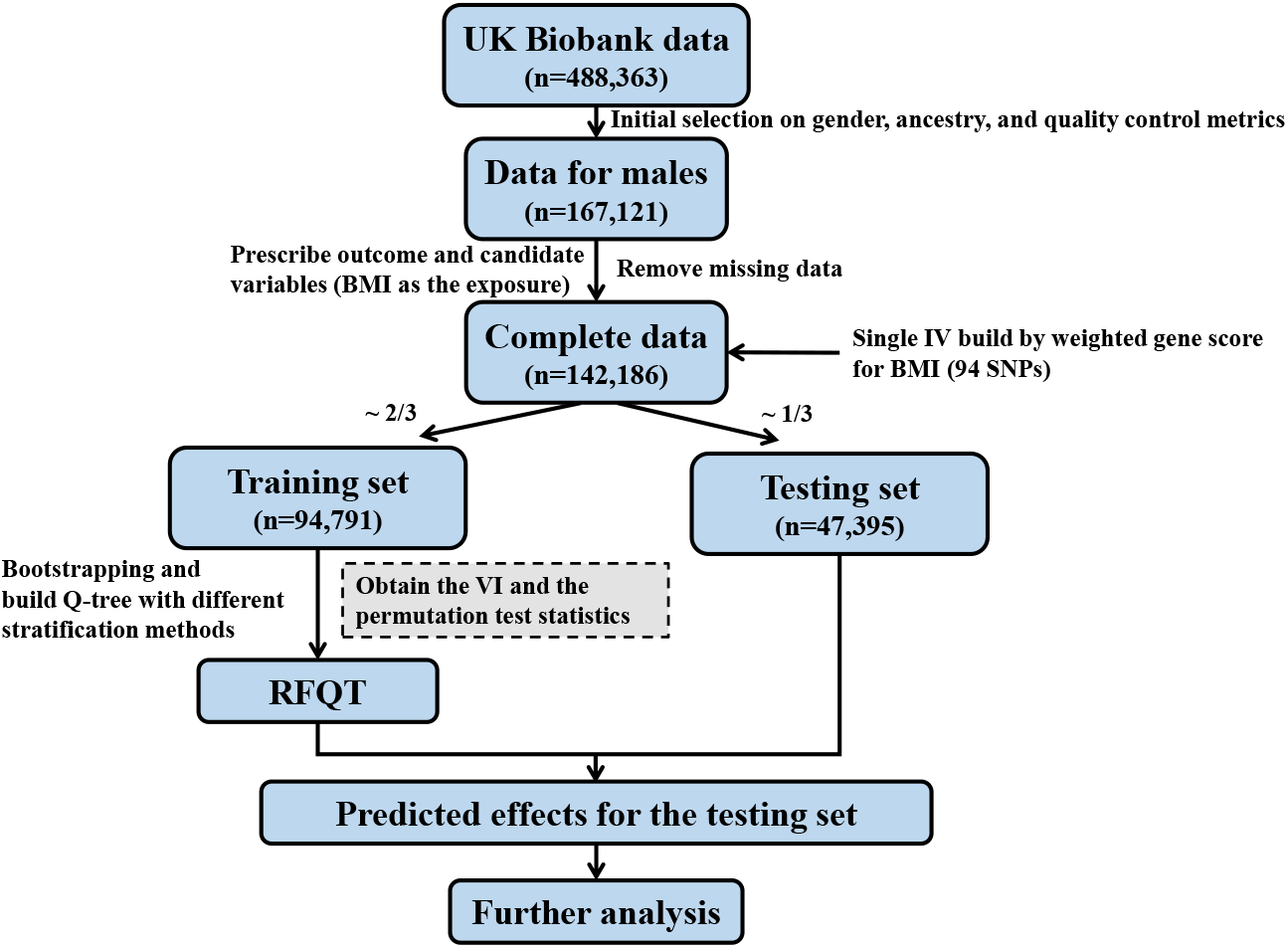
Diagram demonstrating analysis flow for the UK Biobank data. IV: instrumental variable. RFQT: random forest of Q trees. VI: variable importance.

